# Surveillance of COVID-19 vaccine effectiveness – a real-time case-control study in southern Sweden

**DOI:** 10.1101/2021.12.09.21267515

**Authors:** Jonas Björk, Carl Bonander, Mahnaz Moghaddassi, Magnus Rasmussen, Ulf Malmqvist, Fredrik Kahn, Malin Inghammar

## Abstract

The extensive register infrastructure available for COVID-19 surveillance in Scania county, Sweden, makes it possible to classify cases with respect to hospitalization and disease severity, stratify on time since last dose and demographic factors, account for prior infection, and extract data for population controls automatically. Estimated vaccine effectiveness 0-3 months after the last dose remained stable during the study period but waned markedly 6 months after the last dose in older persons.

---

The emergence of new SARS-CoV-2 variants has stressed the importance of continuously surveil COVID-19 vaccine effectiveness and waning immunity (1-4). The population and health care registers in Sweden and the other Nordic countries contain extensive individual-level data for all residents that can be cross-referenced (5). Such register infrastructures offer excellent opportunities for detailed epidemiologic surveillance but are currently underused. The present study aimed to investigate how the Swedish register infrastructure can be used to surveil COVID-19 vaccine effectiveness in real time. To this end, we developed an automatic case-control sampling design using data with complete population coverage from Scania (Skåne), an ethnically and socioeconomically diverse region exceeding 1.3 million inhabitants.

## Surveillance of COVID-19 cases and vaccination

The study cohort included all persons residing in Skåne, southern Sweden, on December 27^th^, 2020 (baseline) when vaccinations started (n = 1 384 531) (6), and was followed for 310 days until November 2^nd^, 2021. Individuals who died or moved out from the region were censored on the date of death or relocation.

The first to be vaccinated in Sweden were nursing home residents, their caregivers and frontline health care workers, followed by the general population in age groups in descending order, currently down to age 12. Three different vaccines have been used in Sweden: BNT16b2 mRNA (Pfizer-BioNTech), mRNA-1273 (Moderna) and ChAdOx1-SARS-CoV-2 (AstraZeneca). The timing of the second dose depended on vaccine type and schedule but was given in median 42 days after the first. From September 1^st^, 2021, a third booster dose was offered, starting with nursing home residents, older people and immunocompromised.

The different data sources were linked using the personal identification number assigned to all Swedish residents (7). Individual-level data on country of birth, civil status, residency and vital status were obtained from the Swedish Total Population Register. Weekly updates on vaccination date, type of vaccine and dose were obtained from the National Vaccination Register, and data on positive SARS-CoV-2 test results from the electronic system SMINet, both kept at the Public Health Agency of Sweden. Data from regional registers and electronic health records were accessed continuously. Hospitalizations 5 days before until 14 days after a positive SARS-CoV-2 test result and U07.1 (ICD-10) among the diagnoses were regarded as caused by COVID-19. Severe disease among the hospitalized was defined as a need of oxygen supply ≥ 5 L/min or admittance to an intensive care unit (ICU).

## Continuous case-control sampling

To avoid conflation of varying infection pressure over time in the population with waning vaccine effectiveness, we used continuous density case-control sampling (8) nested within the study cohort described above. A case was defined as a person with a first-time positive test or any positive test at least 90 days after a prior positive test. For each case, 10 controls without a positive test the same week as the case or 90 days prior were randomly selected from the cohort, matched with respect to sex and age (five-year groups).

## Descriptive results

In total, 96 801 COVID-19 cases occurred during follow-up, of whom 3 334 (3.4%) were hospitalized and 1 303 (1.3%) classified as severe (oxygen supply ≥ 5 L/min or ICU admittance). Identified cases were considerably younger on average towards the end vs. at the start of follow-up (Table 1). BNT16b2 mRNA (Pfizer-BioNTech) was the dominant vaccine type with 79% of all administrated doses.

**Table 1.** Characteristics of the COVID-19 cases and sex and age matched controls, stratified by follow up period

|  | Follow up period |  |  |  |  |  |
| --- | --- | --- | --- | --- | --- | --- |
|  | Period 1, week 10 – 20<br>(day 71 – 147) |  | Period 2, week 21 – 32<br>(day 148 – 231) |  | Period 3, week 33 – 44<br>(day 232 – 310) |  |
|  | Cases | Controls | Cases | Controls | Cases | Controls |
| N | 34 125 | 341 250 | 5 300 | 53 000 | 6 623 | 66 230 |
| Age |  |  |  |  |  |  |
| 0-17 | 18.3 | 18.6 | 19.4 | 21.3 | 31.0 | 29.8 |
| 18-39 | 36.9 | 36.5 | 49.0 | 47.1 | 34.5 | 35.6 |
| 40-64 | 38.2 | 38.2 | 27.9 | 27.9 | 28.2 | 28.2 |
| 65- | 6.7 | 6.7 | 3.7 | 3.7 | 6.4 | 6.4 |
| Sex |  |  |  |  |  |  |
| Females | 50.0 | 50.0 | 50.9 | 50.9 | 52.3 | 52.3 |
| Males | 50.0 | 50.0 | 49.1 | 49.1 | 47.7 | 47.7 |
| Born abroad | 26.8 | 26.4 | 22.0 | 25.9 | 25.0 | 25.6 |
| Civil status |  |  |  |  |  |  |
| Married | 39.5 | 35.9 | 35.4 | 32.0 | 28.3 | 25.0 |
| Widow/widower | 2.6 | 2.6 | 1.2 | 1.2 | 0.8 | 0.8 |
| Divorced | 10.3 | 10.4 | 8.4 | 8.9 | 6.8 | 6.6 |
| Single | 47.6 | 51.1 | 55.0 | 57.9 | 64.0 | 67.6 |
| Vaccine doses |  |  |  |  |  |  |
| 0 | 94.9 | 91.0 | 78.1 | 70.2 | 64.6 | 44.9 |
| 1 | 4.2 | 5.6 | 15.5 | 18.0 | 8.9 | 11.3 |
| 2 | 0.9 | 3.4 | 6.3 | 11.8 | 26.4 | 43.7 |
| 3 |  |  |  |  | 0.1 | 0.1 |
| Vaccine type, at least two doses | N=302 | N=11 497 | N=335 | N=6 246 | N=1 757 | N=30 758 |
| Pfizer | 91.7 | 90.5 | 71.0 | 72.0 | 79.3 | 81.4 |
| Moderna | 6.6 | 8.2 | 10.7 | 11.5 | 13.0 | 6.0 |
| AZ | 1.3 | 0.7 | 12.2 | 8.7 | 4.9 | 8.8 |
| Mixed | 0.3 | 0.6 | 6.0 | 7.7 | 2.8 | 3.8 |
| Time since last dose |  |  |  |  |  |  |
| 0 – 3 months | 87.7 | 92.0 | 69.3 | 77.8 | 52.5 | 68.2 |
| 3 – 6 months | 12.3 | 8.0 | 28.4 | 21.1 | 33.8 | 25.7 |
| ≥ 6 months | 0.0 | 0.0 | 2.4 | 1.2 | 13.7 | 6.1 |
| Prior SARS-CoV-2 infection | 0.6 | 6.7 | 2.7 | 10.3 | 2.6 | 11.8 |
| Hospitalized | 3.1 | - | 2.0 | - | 3.0 | - |

## Estimating vaccine effectiveness

We used conditional logistic regression (Stata SE 14.2, Stata Corp) for the 1:10 case : control matched sets to estimate the odds ratio (OR) and vaccine effectiveness (VE = 1 – OR) together with 95% confidence interval (CI, accounting for individual clustering in the full-period estimates) for the association between vaccination status and risk of infection (positive test), being hospitalized or developing severe COVID-19. Only doses received at least 7 days before the case date were counted within each matched set when vaccination status was assessed. Vaccination status among those with at least two doses was grouped according to time since last dose (0 – 3, 3 – 6 or > 6 months) and vaccine type. Results were (besides the matching) presented unadjusted, but prior infection was included in a sensitivity analysis.

## Surveillance results

Vaccine effectiveness was monitored weekly, from week 10 (March 8^th^, 70 days after vaccination start) to week 44 (November 2^nd^, 310 days after vaccination start). The estimated effectiveness against infection after two doses of any of three vaccines was 76% in median, and the weekly estimates varied between 48% and 88% with no evident time trend (Figure 1). Two doses of any of the three vaccines offered high protection against hospitalization (median effectiveness 87%, range 71-95%) and severe disease (median effectiveness 92%, range 74-100%), here aggregated monthly due to small numbers (Figure 2). Estimated effectiveness against infection (Supplementary Figure 1A) and hospitalization (Supplementary Figure 1B) 0-3 months after the last dose remained stable during the study period. Waning effectiveness was consistently noted 6 months after the last dose, but with substantial fluctuations due to the small number of hospitalized.

**Figure 1.**
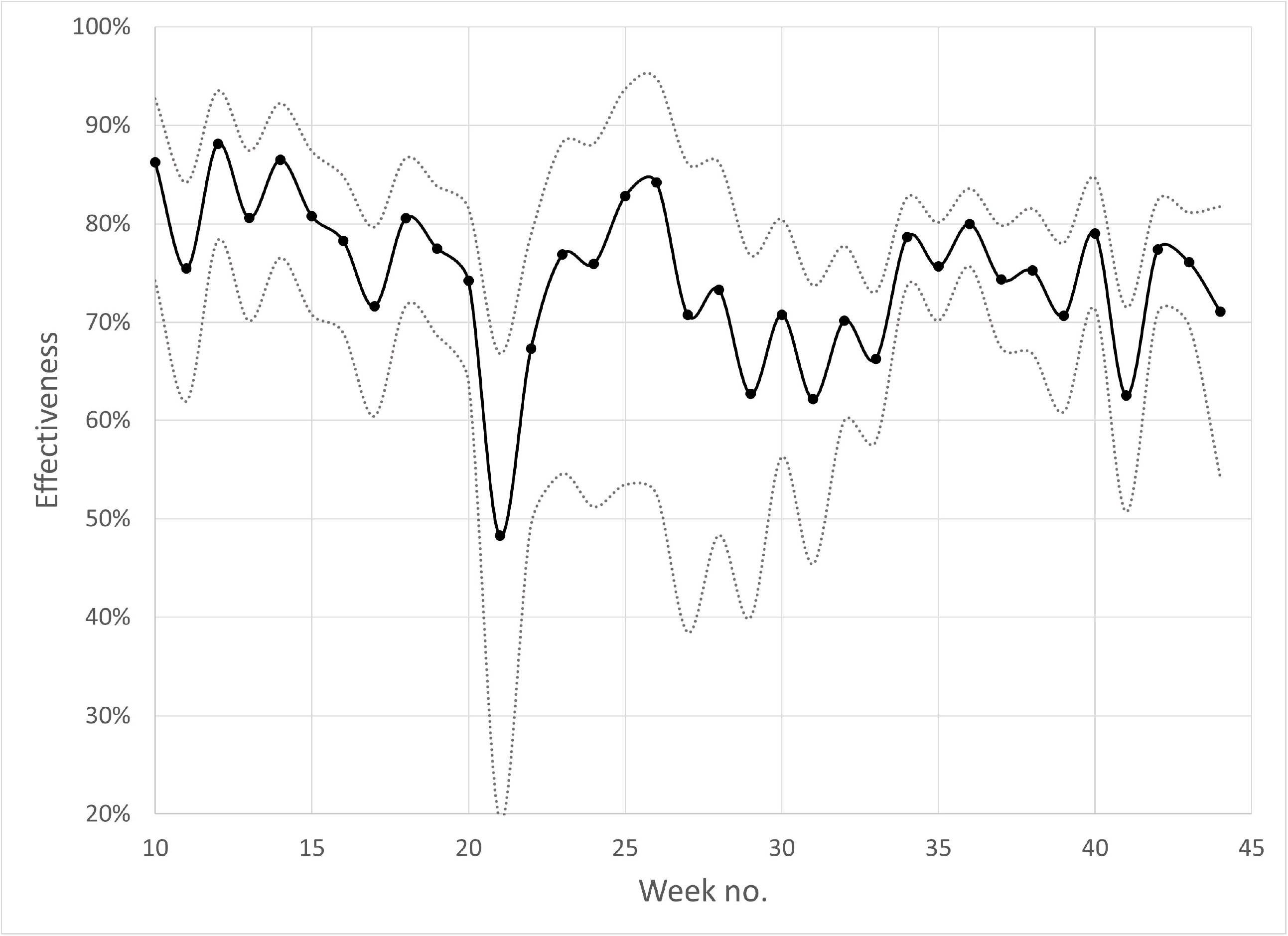
Weekly surveillance in Scania county, Southern Sweden, during week 2020 10 – 44 of the estimated effectiveness against SARS-CoV-2 infection after two doses of any of three vaccines. Grey dotted lines represent 95% confidence intervals.

**Figure 2.**
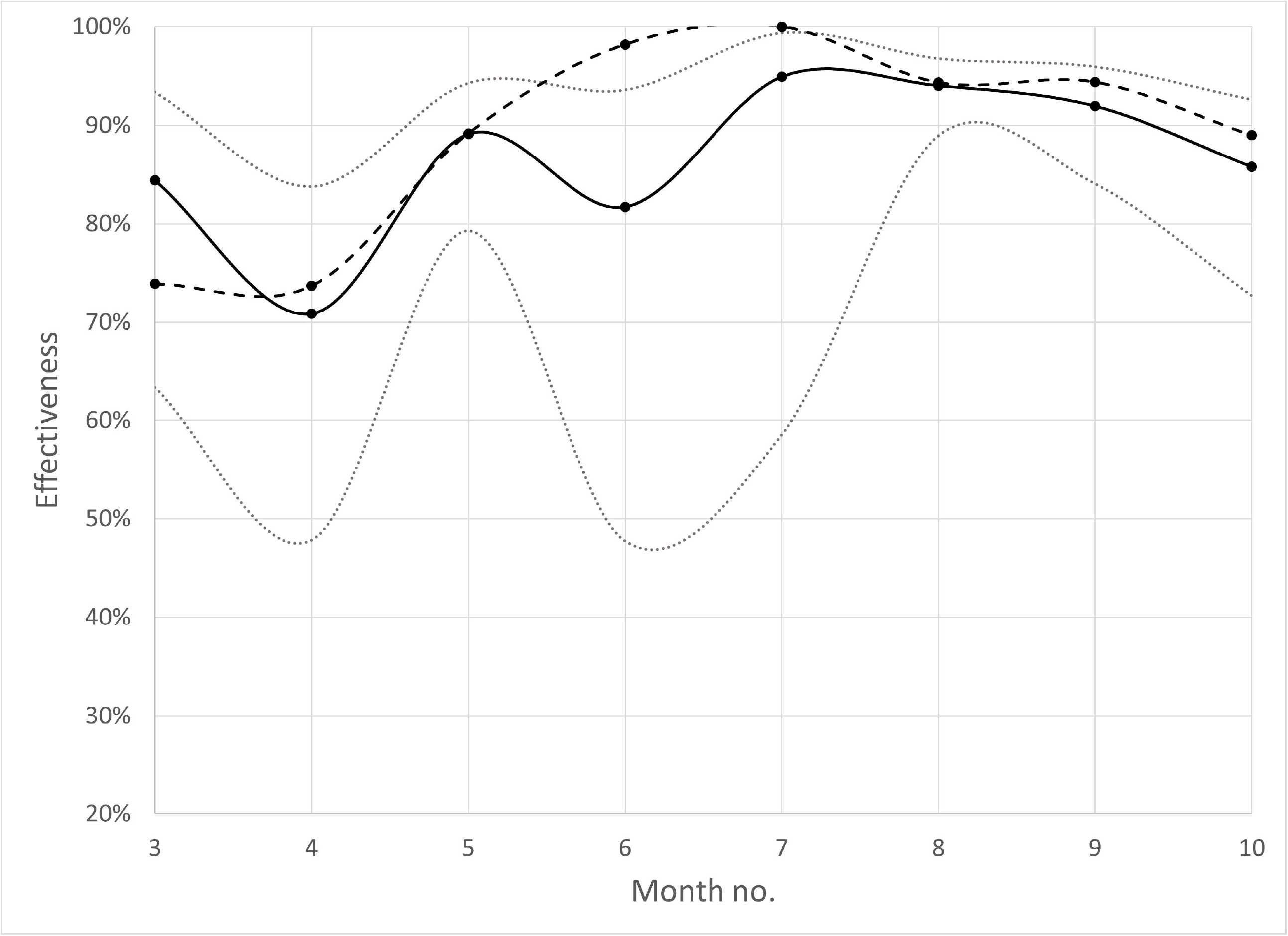
Monthly surveillance in Scania county, Southern Sweden, during March - October 2020 of the estimated effectiveness against COVID-19 hospitalization (solid black curve) and severe disease (oxygen supply ≥ 5 L/min or ICU admittance; dotted black curve) after two doses of any of three vaccines. Grey dotted lines represent 95% confidence intervals for effectiveness against hospitalization.

### Average effectiveness against infection, hospitalization and severe disease

The mRNA vaccines (Pfizer-BioNTech and Moderna) exhibited higher effectiveness on average against infection than the vector vaccine (AstraZeneca; Figure 3). All three vaccines offered strong protection against hospitalization and severe disease in individuals ≥ 65 years (Supplementary Figure 2). Lower effectiveness against hospitalization and severe disease was indicated for mRNA-1273 (Moderna) in individuals below 65 years. Vaccine effectiveness waned considerably with time since last dose, especially in individuals ≥ 65 years (Figure 3 and Supplementary Figure 2). The protection against hospitalization and severe disease remained more satisfactory in younger individuals but was also more statistically uncertain. Prior SARS-CoV-2 infection offered strong protection against new infection (average effectiveness 86%, 95% CI 85-87%) and hospitalization (average effectiveness 91%, 95% CI 82-96%, not in tables), but did not confound the estimates of vaccine effectiveness.

**Figure 3.**
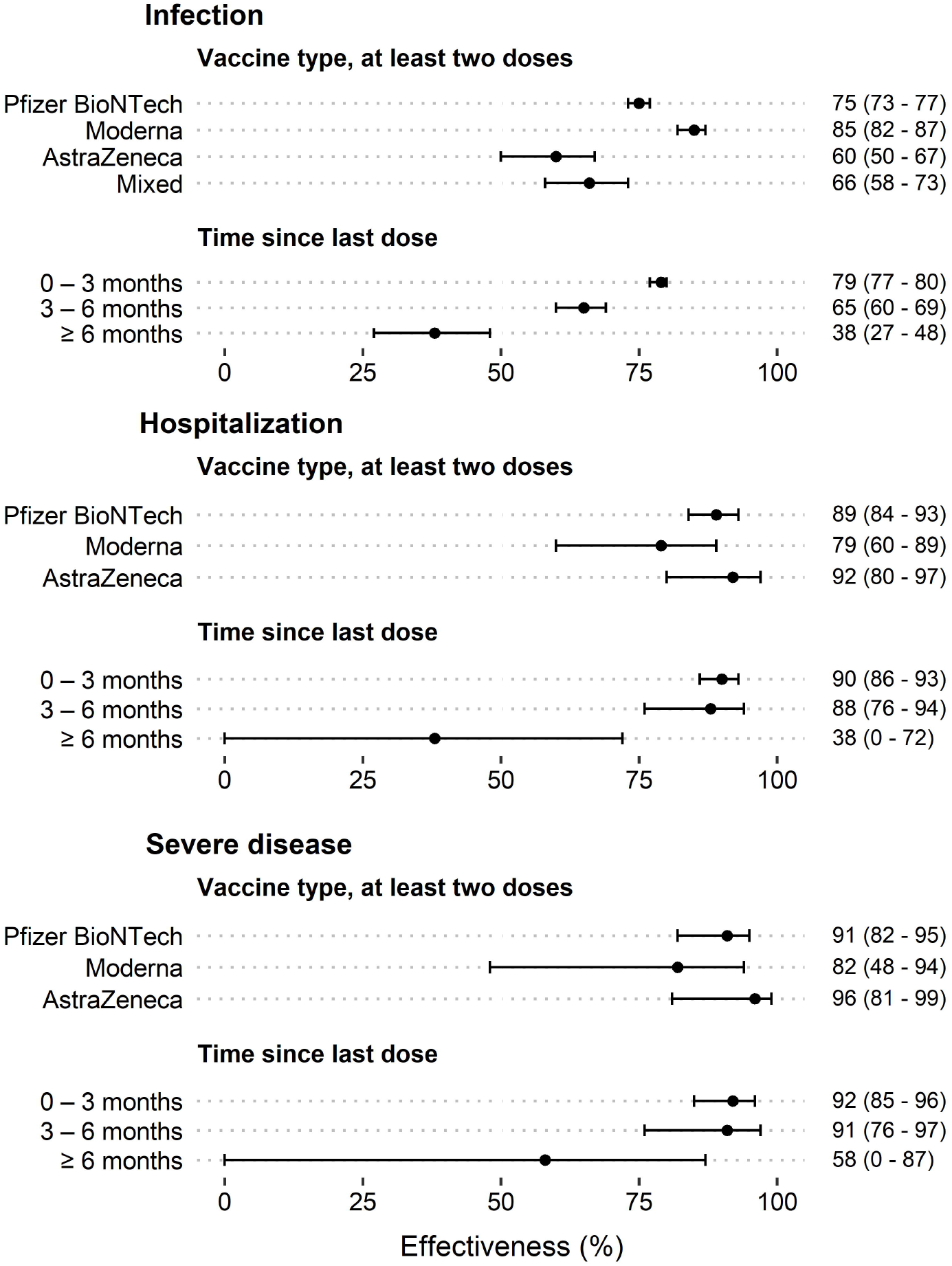
Average effectiveness (percent; 95% cluster-robust CIs) of the COVID-19 vaccination in protecting infection, hospitalization and severe disease (oxygen supply ≥ 5 L/min or ICU admittance) in relation to vaccine type and time since last dose.

## Discussion

The extensive register data available in our real-time surveillance system allows us to classify hospitalized cases further concerning disease severity, stratify by time since last dose and demographic factors, account for prior SARS-CoV-2 infection and extract data for population controls automatically. The density case-control sampling is a convenient approach to avoid bias from underlying time trends in the population (8). Such bias could occur if, e.g., associations between age or time since last dose among the vaccinated and the infection pressure on society are not considered during design or analysis. Selective COVID-19 testing depending on vaccination status could still bias the case-control sample but is unlikely to impact estimated effectiveness against hospitalization and severe disease.

The surveillance was hampered by limited population size and a low infection rate during parts of the study period. As a comparison, the population in Scania county is only 1/7 of the Israeli population, a well-known example where continuous surveillance of COVID-19 vaccine effectiveness is conducted (3). However, after adaptions, our extensive surveillance system should be possible to implement on the national level in Sweden, with a population exceeding 10M.

## Conclusion

The present investigation demonstrates the strength of combining individual-level population and health care register data to monitor vaccine effectiveness in real time. A natural extension of the surveillance system would be to add further individual-level data on socioeconomic conditions, disease histories and care needs among older people, all available in the Swedish register infrastructure. Such additions would make it possible to monitor protection in vulnerable populations separately as a basis for decisions on additional booster doses, campaigns to increase vaccine uptakes in subgroups or other directed interventions.

## Data Availability

All data produced in the present study are available upon reasonable request to the authors, and given that relevant permissions are obtained.

## Ethics and Permissions

Ethical approval was obtained from the Swedish Ethical Review Authority (2021-00059).

