## Supplementary material for "Surveillance of COVID-19 vaccine effectiveness – a real-time case-control study in southern Sweden"

**Supplementary Figure 1.** Monthly surveillance in Scania county, Southern Sweden, during March - October 2020 of the estimated effectiveness against **A)** SARS-CoV-2 infection, **B)** COVID-19 hospitalization. Solid curves represent 0-3, dotted curves 3-6 and dashed curves more than 6 months since the last dose of any of three vaccines.

**A.**

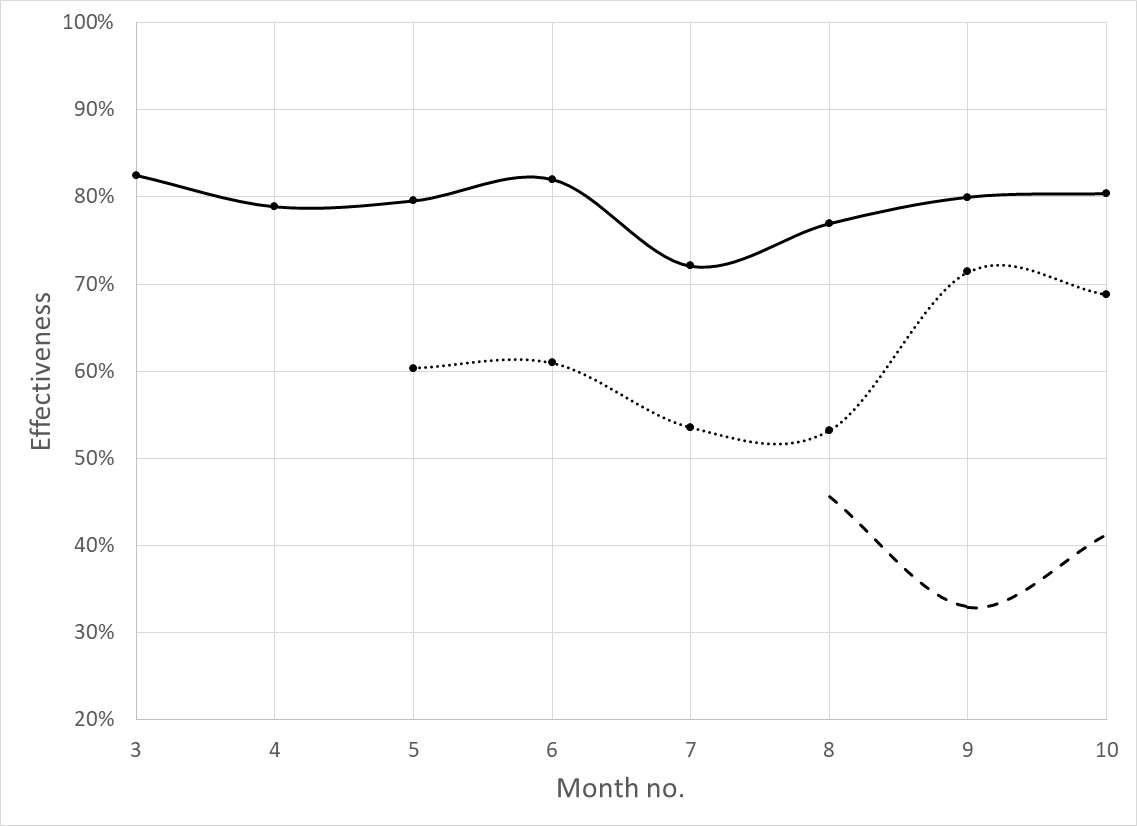

**B.**

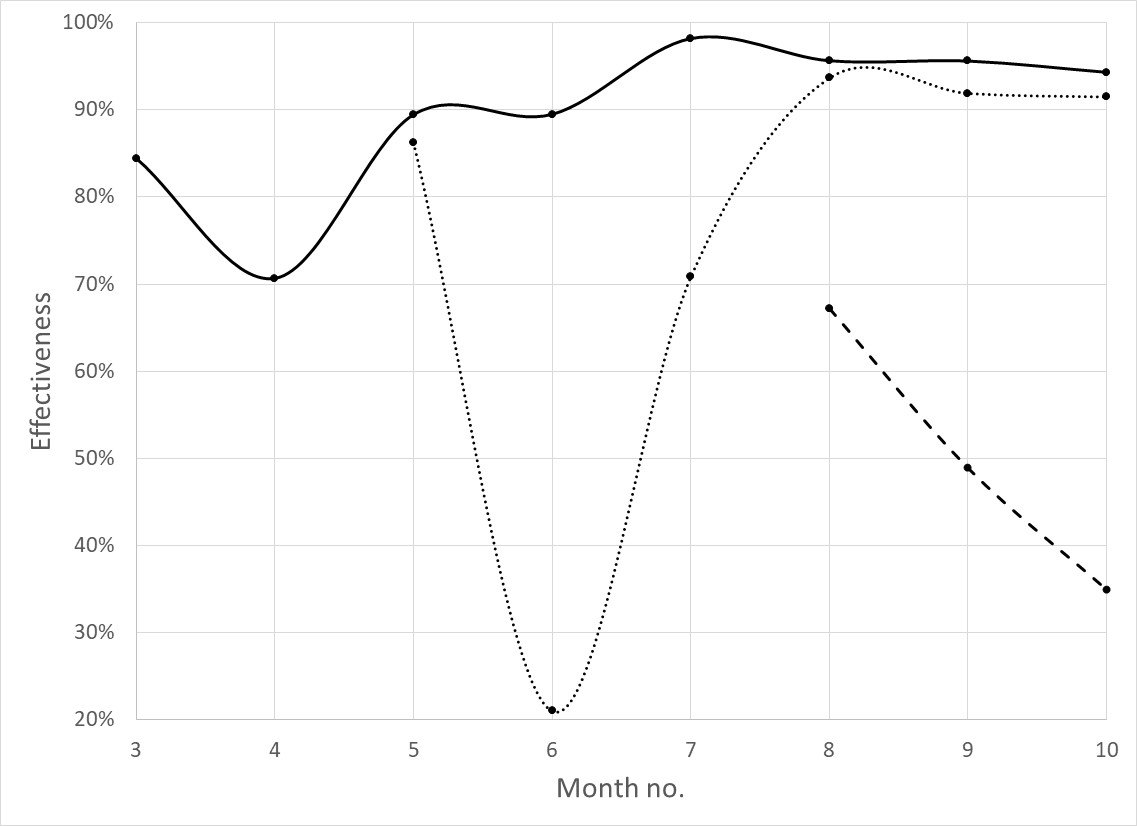

**Supplementary Figure 2.** Average effectiveness (percent; 95% cluster-robust CIs) of the COVID-19 vaccination in protecting infection, hospitalization and severe disease (oxygen supply ≥ 5 L/min or ICU admittance) in relation to vaccine type and time since last dose, stratified by age.

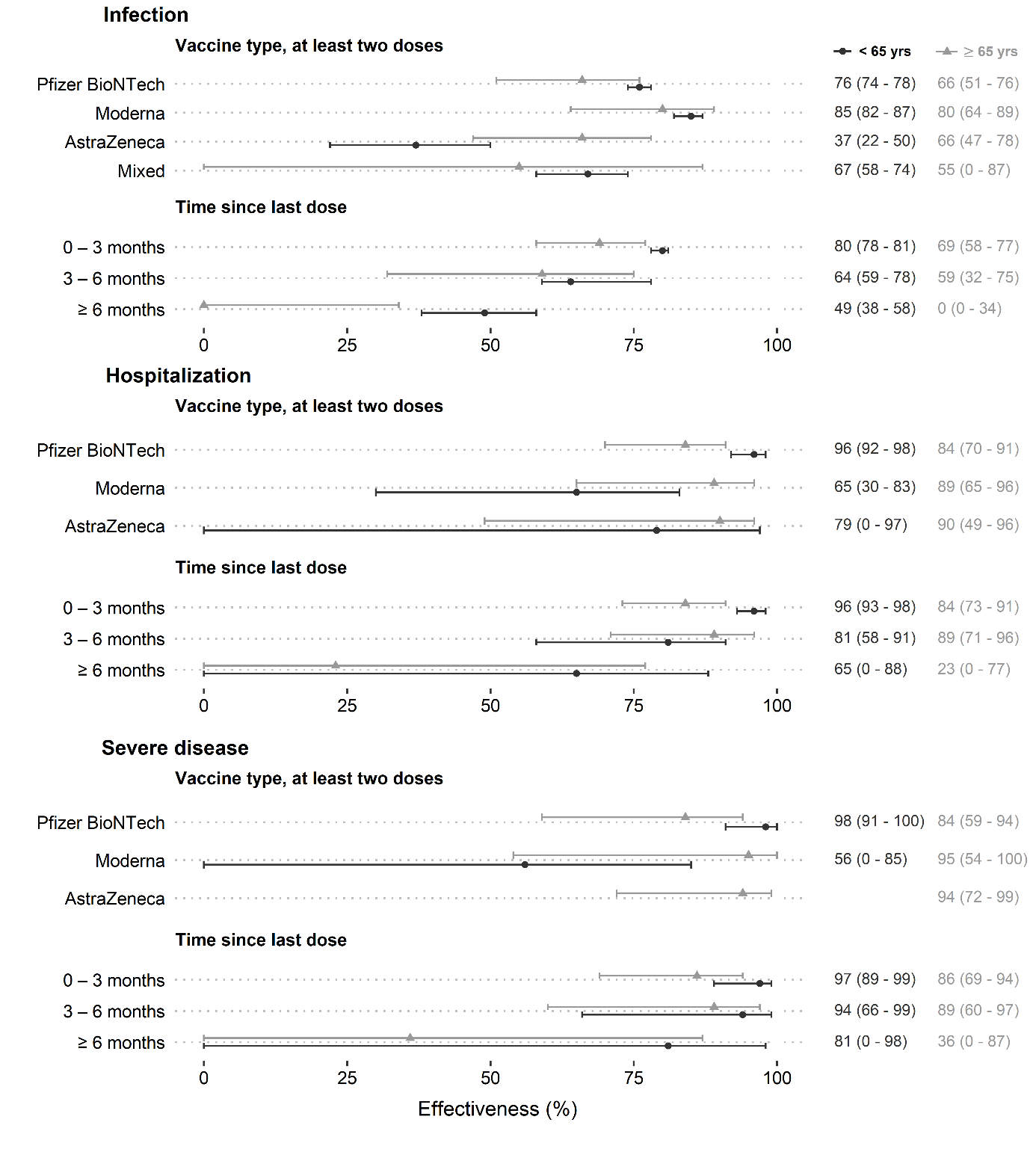
